# Metabolic Insights into Iron Deposition in Relapsing-Remitting Multiple Sclerosis via 7T Magnetic Resonance Spectroscopic Imaging

**DOI:** 10.1101/2023.03.28.23287856

**Authors:** Alexandra Lipka, Wolfgang Bogner, Assunta Dal-Bianco, Gilbert J. Hangel, Paulus S. Rommer, Bernhard Strasser, Stanislav Motyka, Lukas Hingerl, Thomas Berger, Fritz Leutmezer, Stephan Gruber, Siegfried Trattnig, Eva Niess

**Author notes:** **Corresponding author:** Wolfgang Bogner; High Field MR Centre, Department of Biomedical Imaging and Image-guided Therapy, Medical University of Vienna, Lazarettgasse 14, A-1090 Vienna, Austria. **Institutional address:** High Field MR Centre, Department of Biomedical Imaging and Image-guided Therapy, Medical University of Vienna, Lazarettgasse 14, A-1090 Vienna, Austria.

## Abstract

**Objective:** To investigate the metabolic pattern of different types of iron accumulation in multiple sclerosis (MS) lesions, and compare metabolic alterations within and at the periphery of lesions and newly emerging lesions *in vivo* according to iron deposition.

**Methods:** 7T MR spectroscopic imaging and susceptibility-weighted imaging was performed in 31 patients with relapsing-remitting MS (16 female/15 male; mean age, 36.9 ± 10.3 years). Mean metabolic ratios of four neuro-metabolites were calculated for regions of interest (ROI) of normal appearing white matter (NAWM), “non-iron” (lesion without iron accumulation on SWI), and three distinct types of iron-laden lesions (“rim”: distinct rim-shaped iron accumulation; “area”: iron deposition across the entire lesions; “transition”: transition between “area” and “rim” accumulation shape), and for lesion layers of “non-iron” and “rim” lesions. Furthermore, newly emerging “non-iron” and “iron” lesions were compared longitudinally, as measured before their appearance and one year later.

**Results:** Thirty-nine of 75 iron-containing lesions showed no distinct paramagnetic rim. Of these, “area” lesions exhibited a 65% higher mIns/tNAA (p=0.035) than “rim” lesions. Comparing lesion layers of both “non-iron” and “rim” lesions, a steeper metabolic gradient of mIns/tNAA (“non-iron” +15%, “rim” +40%) and tNAA/tCr (“non-iron” −15%, “rim” −35%) was found in “iron” lesions, with the lesion core showing +22% higher mIns/tNAA (p=0.005) and −23% lower tNAA/tCr (p=0.048) in “iron” compared to “non-iron” lesions. In newly emerging lesions, 18 of 39 showed iron accumulation, with the drop in tNAA/tCr after lesion formation remaining significantly lower compared to pre-lesional tissue over time in “iron” lesions (year 0: p=0.013, year 1: p=0.041) as opposed to “non-iron” lesions (year 0: p=0.022, year 1: p=0.231).

**Conclusion:** 7T MRSI allows ***in vivo*** characterization of different iron accumulation types each presenting with a distinct metabolic profile. Furthermore, the larger extent of neuronal damage in lesions with a distinct iron rim was reconfirmed via reduced tNAA/tCr concentrations, but with metabolic differences in lesion development between (non)-iron-containing lesions. This highlights the ability of MRSI to further investigate different types of iron accumulation and suggests possible implications for disease monitoring.

**Key points:**

- Iron-containing lesions were suggested as a biomarker for tissue damage, a more aggressive disease course, and worse clinical outcome, but related metabolic alterations are poorly understood.
- Our MRSI results confirm a higher extent of tissue damage within paramagnetic rim lesions reflected by reduced tNAA/tCr. Forty-six percent of newly emerging lesions showed an iron accumulation, correlating with an altered metabolic behavior compared to non-iron lesions.
- Only forty-eight percent of iron-containing lesions have a distinct rim-shaped iron accumulation, although most studies focus on these paramagnetic rim lesions. Our results show highly different metabolic profiles (especially with regard to mIns/tNAA and tNAA/tCr) within different iron accumulation types, highlighting the need for distinct classification of iron accumulation in future studies.

## 1. Introduction

T1- and T2-weighted magnetic resonance imaging (MRI) is an integral part of diagnosis and treatment monitoring in Multiple Sclerosis (MS) (Polman et al., 2011; Thompson et al., 2018; Filippi et al., 2022). Yet, the biggest drawback of MRI measures is the inability to fully explain the clinical status (McFarland, 1999). Currently established MRI techniques are not specific enough to assess the underlying pathological process, as they are sensitive only to macroscopic tissue damage. MR Spectroscopic Imaging (MRSI), however, can visualize pathology on a biochemical level by mapping the spatial distribution of various brain metabolites (Öz et al., 2014; Hangel et al., 2022). The most commonly reported abnormalities found in MS are decreased N-acetylaspartate (NAA; reflecting reduced neuronal/axonal integrity and function) and increased myo-inositol (mIns; a marker for astroglial hypertrophy and hyperplasia), which correlates with clinical impairment, and elevated choline (Cho; a marker of myelin turnover) (De Stefano et al., 2002; Filippi et al., 2003; Kirov et al., 2009). Susceptibility Weighted Imaging (SWI) can provide additional information about tissue microstructure and, specifically, iron deposition. Iron rim lesions, a subset of chronically active lesions, which are usually defined as having a demyelinated core surrounded by a rim of reactive astrocytes and iron-laden microglia/macrophages (Martire et al., 2022), can be detected as hypointense paramagnetic rim lesions (PRLs) on SWI. These iron-rim lesions present early in lesion develpment as a contrast-enhanced active lesion with a phase rim that persists in its chronic stage. In contrast to lesions with only transient phase rim, they show less shrinkage and have been linked to tissue damage, a more progressive disease course, and, thus, a worse clinical outcome (Fischer et al., 2013; Absinta et al., 2016; Hametner et al., 2018; Luchetti et al., 2018; Absinta et al., 2019). Iron rims are observed to diminish over time, and the lesion may transition to an inactive stage and may eventually remyelinate (Dal-Bianco et al., 2017, 2021).

A recent review article (Martire et al., 2022) has suggested iron-containing lesions as a new lesion biomarker for MS (Kolb et al., 2022), as those lesions can be found in a high proportion of MS patients regardless of their clinical phenotype (Absinta et al., 2016; Dal-Bianco et al., 2017; Absinta et al., 2019; Kaunzner et al., 2019). Furthermore, the high specificity of this technique to differentiate MS from MS-mimicking diseases (Maggi et al., 2018; Filippi et al., 2019) and the potential to predict the conversion from radiologically isolated syndrome (Lim et al., 2022) and clinically isolated syndrome (Clarke et al., 2020) has been shown.

Iron-containing lesions have mostly been studied regardless of their iron deposition subtype (Dal-Bianco et al., 2017, 2021) and their characteristic slow expansion is currently controversially discussed (Absinta & Dal-Bianco, 2021; Arnold et al., 2021; Enzinger, 2021). 7T MRI can, due to its higher spatial resolution, differentiate between iron-containing lesions with a distinct paramagnetic rim (herein “rim”) and those with a diffuse iron accumulation that involves the whole lesion(herein “area”) (Hametner et al., 2018). Furthermore, the higher spatial resolution at 7T enables the study of metabolic alterations during lesion development (Lipka et al., 2023).

Metabolic alterations as measured by MRSI haveLto the best of our knowledgeLnever been investigated within different types of iron-accumulating lesions. Therefore, we aim to: (1) characterize different types of iron-containing lesions and determine the differences in their metabolic alterations; (2) investigate differences in metabolic alterations inside and at the periphery of iron- and non-iron-containing lesions; and (3) identify metabolic differences in newly emerging iron- and non-iron-containing lesions.

## 2. Materials and Methods

### 2.1. Study Population

For this prospective study, written, informed consent and IRB approval (EK 154/2009) were obtained. Patient recruitment by the Department of Neurology took place between January 2016 and December 2017 and fulfilled the following criteria: clinically definite MS diagnosis according to revised McDonald criteria (Polman et al., 2011); no change in the Expanded Disability Status Scale (EDSS) score (Kurtzke, 2015) during the prior six months; stable/no treatment during the prior six months; no relapse or corticosteroid treatment within the prior three months; no other neuropsychiatric or neurological disease; and no 7T-MRI contraindications (Figure 1). Detailed descriptions on overlapping study populations with Dal-Bianco et al. and Lipka et al. (Dal-Bianco et al., 2021; Lipka et al., 2023) can be found in Suppl. Text 1.

**Figure 1:**
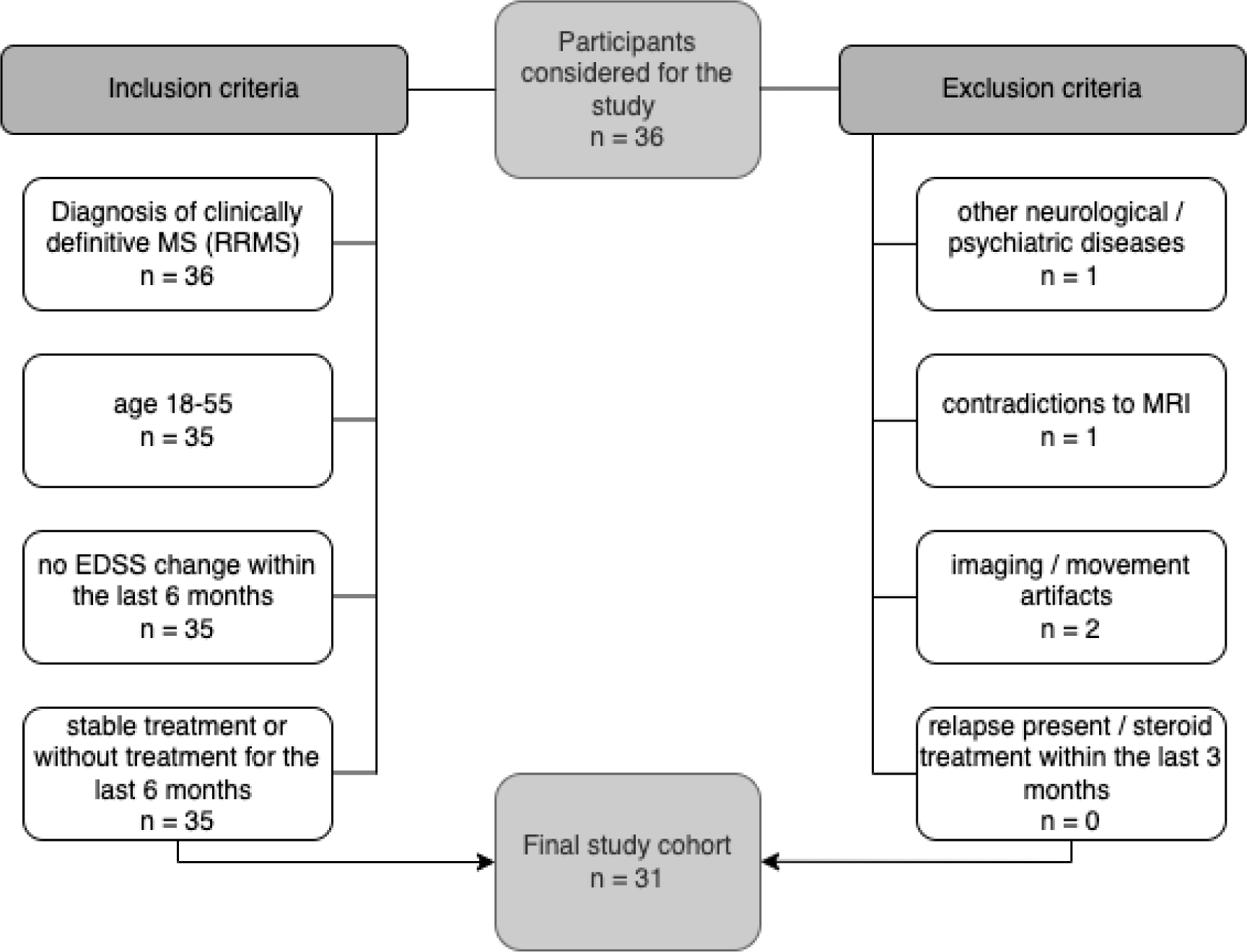
Flowchart of participants with MS enrolled in the study. EDSS = Expanded Disability Status Scale; MS = multiple sclerosis.

### 2.2. Imaging Protocol

All measurements were performed on the same 7T whole-body MR scanner (Magnetom, Siemens Healthcare, Erlangen, Germany) using a 1TX/32RX head coil (Nova Medical, Wilmington, MA). Each session included three-dimensional T_1_-weighted imaging using two magnetization-prepared rapid gradient echoes (MP2RAGE) (TR=5000ms, TE=4.13ms, TI1/TI2=700ms/2700ms, scan time=8:02min) with 0.8×0.8×0.8mm^3^ spatial resolution, three-dimensional T_2_-weighted imaging using fluid-attenuated inversion recovery (FLAIR) (TR=8000ms, TE=270ms, TI=2180ms, scan time=7:14min) with 0.86×0.86×0.86 mm^3^ spatial resolution, and three-dimensional susceptibility weighted imaging (SWI) (TR=38ms, TE=25ms, scan time=7:24min) with 0.3×0.3×1.2mm^3^ spatial resolution to visualize MS lesions and potential iron deposition prior to positioning of the MRSI volume of interest. A single-slice, transversal, two-dimensional free induction decay MRSI (Heckova et al., 2019) scan was obtained above the corpus callosum with an acquisition delay/repetition time of 1.3ms/200ms, an Ernst flip angle of 27°, four-fold parallel imaging acceleration via Controlled Aliasing in Parallel Imaging Results in Higher Acceleration(CAIPIRINHA) (Strasser et al., 2017), a field of view of 220×220mm^2^, a matrix size of 100×100, a nominal voxel size of 2.2×2.2×8mm^3^, an effective voxel volume of 77μl (as explained in Kreis et al (Kreis et al., 2021), nominal voxel volume, 38μl), and a scan time of 6:06min (Hangel et al., 2018; Heckova et al., 2019). Additional data acquisition details can be found in Suppl. Table 1.

### 2.3. Spectroscopic Data Processing

After extracting brain masks from T_1_-weighted images using the FSL brain extraction tool (Smith, 2002), MRSI data from inside the brain were processed with an automated in-house-developed post-processing pipeline using Matlab (R2013a | Mathworks, Natick, MA), MINC (v2.0 | McConnell Brain Imaging Center, Montreal, Quebec, Canada) and Bash (v4.2.25 | Free Software Foundation, Boston, MA). The pipeline included MUSICAL coil combination (Strasser et al., 2013), 2D-CAIPIRINHA reconstruction (Strasser et al., 2017), lipid signal removal via L2-regularization (Bilgic et al., 2014), and spatial Hamming filtering. Using a basis-set of 17 simulated metabolites (Naressi et al., 2001) and a measured macromolecular background (Považan et al., 2015), individual spectra were fitted in the spectral range of 1.8-4.2 ppm with LCModel (version 6.3-1; http://s-provencher.com/lcmodel.shtml). Afterward, metabolic maps and their ratios, quantification precision [i.e., Cramer-Rao Lower Bounds (CRLBs)], spectral quality [i.e., FWHM (linewidth as full-width-half-maximum of the fitted NAA peak), SNR (signal-to-noise ratio of the fitted NAA peak)] were created (Suppl. Table 2). MRS data reporting followed standardized guidelines (Lin et al., 2021) (Suppl. Table 1). For the following analysis of spectroscopic data, metabolic ratio maps were used, as we did not acquire additional water-unsuppressed MRSI.

### 2.4. Data Analysis

#### 2.4.1. Segmentation and iron accumulation type categorization

After resampling (via tricubic interpolation) of metabolic maps to the resolution of T1-weighted images, lesion and representative (fixed volume of 37mm^3^, minimum distance of 0.5cm to GM/CSF/lesions) normal-appearing white matter “NAWM” ROIs were segmented via the user-guided, semi-automatic segmentation software ITK–SNAP (Yushkevich et al., 2006). Based on SWI images, ROIs were qualitatively categorized into four iron accumulation types by two authors individually (A.D.B with 10 years of experience, A.L. with three years of experience) if the iron deposition spanned three contiguous layers: (a) “rim” (distinct rim-shaped iron deposition); (b) “area” (iron accumulation covering the entire lesion); (c) “transition” (transition between “area” to “rim” shape); and (d) “non iron” if no iron accumulation was present (Figure 2). For the final analysis, only lesions exceeding 20mm^3^ were included to mitigate the influence of partial volume errors.

**Figure 2:**
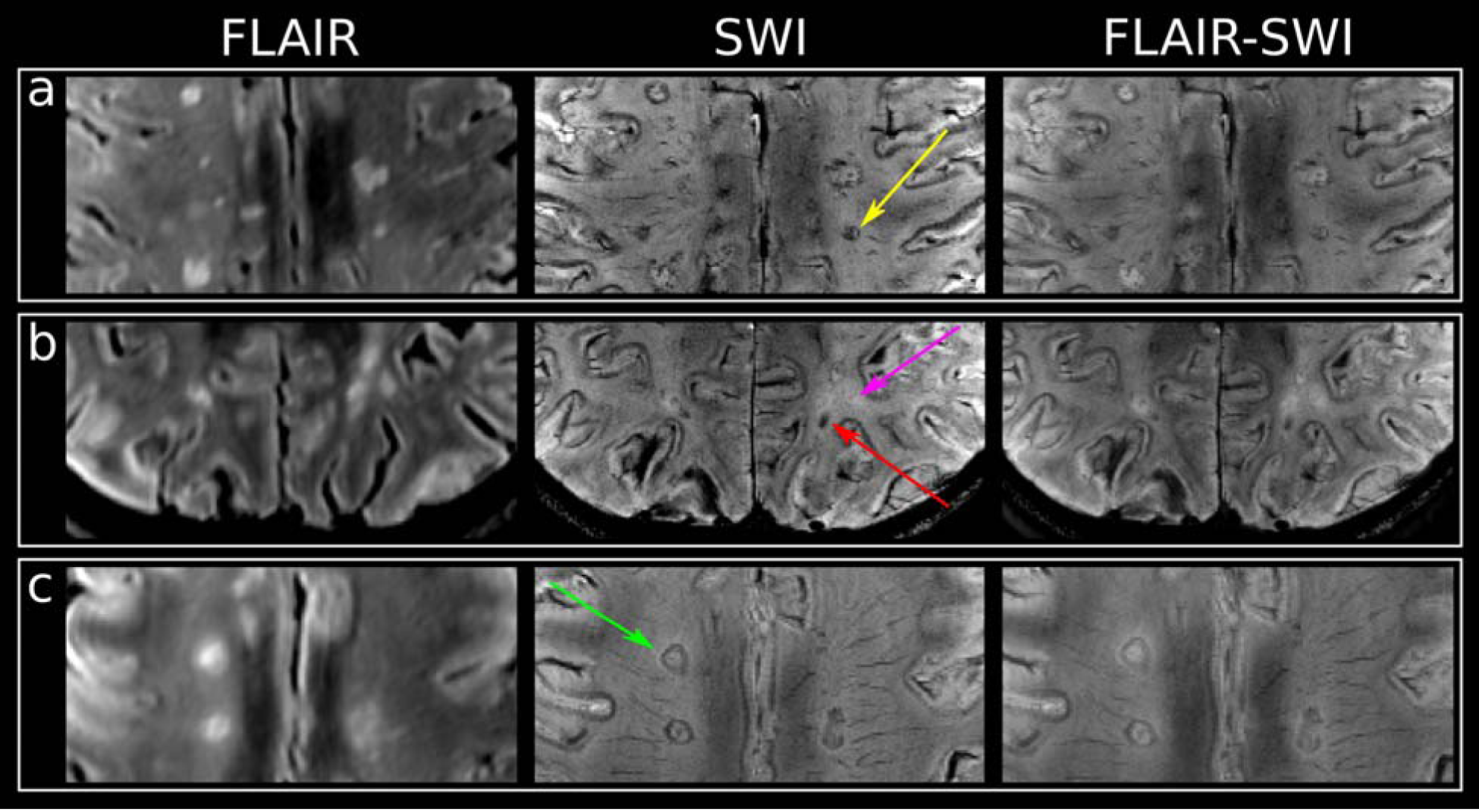
T2-FLAIR, SWI, and overlaid FLAIR-SWI images of (a) lesions categorized as “transition” with yellow arrow pointing toward an example, (b) “area” iron deposition indicated by a red arrow, “non-iron” indicated by a pink arrow, and (c) “rim” lesion with an iron rim indicated by a green arrow.

#### 2.4.2. Layer analysis

To investigate whether the metabolic profile within and in the proximity of MS lesions differed between “non-iron” and “rim” lesions, the segmented ROIs of these categories were both dilated and eroded three times. This resulted in seven lesion layer rings in total, spanning from the outermost layer (L+3) to the innermost layer (L-3), each ∼1mm thick. Voxels that potentially intruded into the GM and CSF after dilation were handled in the following way: lesion-free GM and CSF masks (created using Freesurfer and mincmath) were subtracted from the respective ROIs; where needed, manual correction for GM, CSF and voxels of neighboring lesions was performed using FSLView (Figure 5A). Because at least four lesion layers were required to “fit” inside the lesion, only lesions of an original size of 100-400mm3 were included. For each lesion, metabolic ratios were normalized to NAWM (represented by the respective outermost lesion layer).

**Figure 3:**
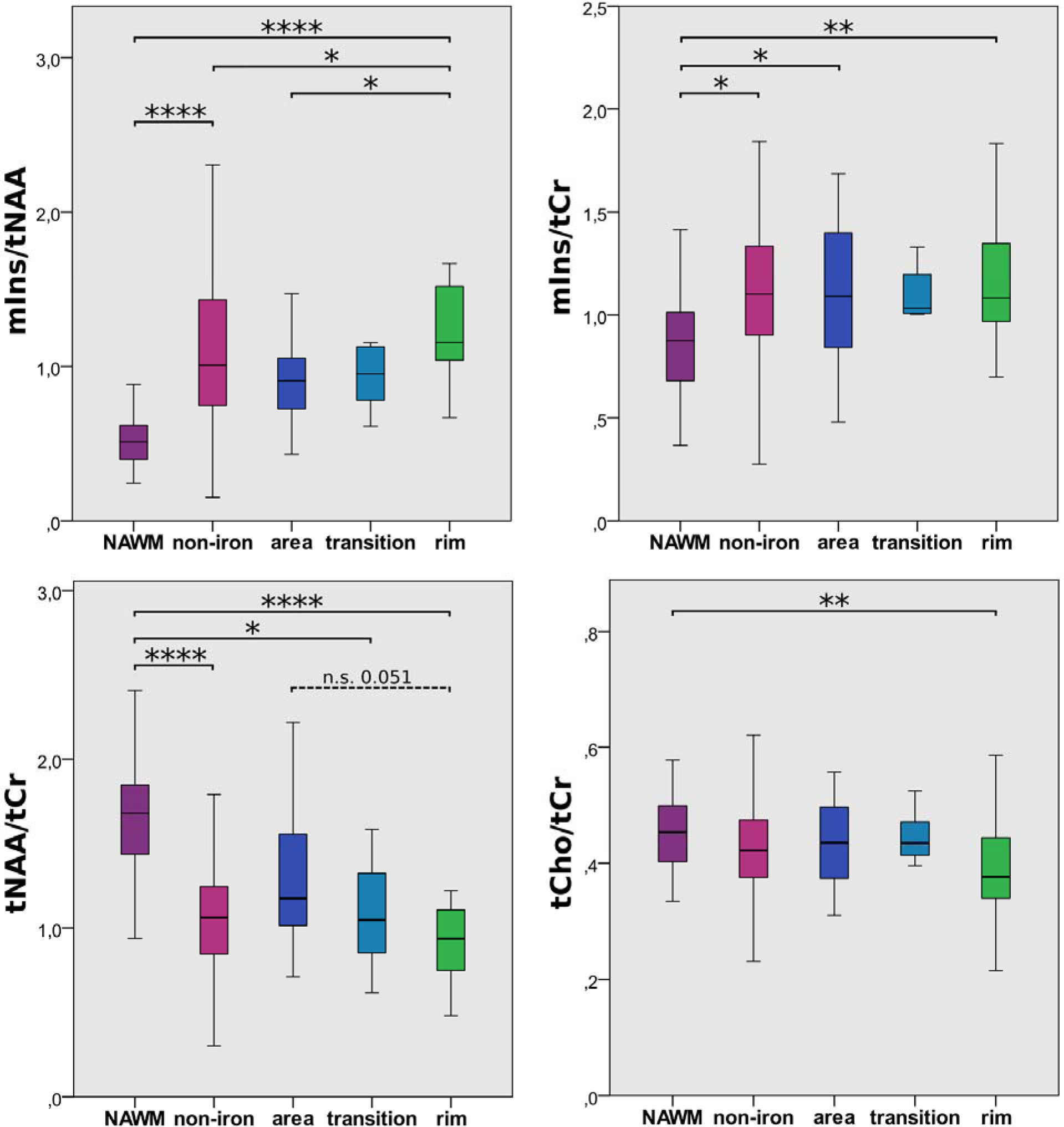
Boxplot diagram for mIns/tNAA, mIns/tCr, tNAA/tCr, and tCho/tCr of the NAWM and the different lesion categories “non-iron,”, “area,” “transition,” and “rim.” Significant differences were found, in particular, for mIns/tNAA and tNAA/tCr.

**Figure 4:**
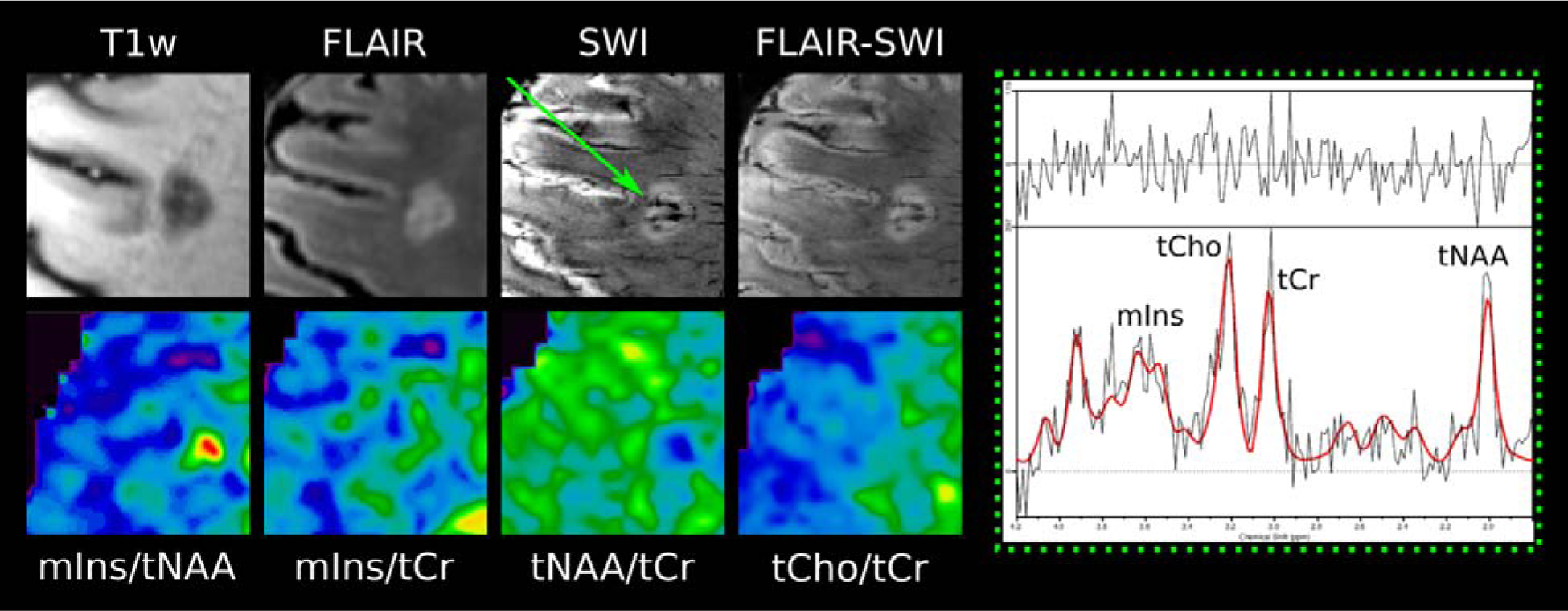
Examples of T1-weighted MP2RAGE, T2-weighted FLAIR, SWI, and overlaid FLAIR-SWI, as well as metabolic maps of mIns/tNAA, mIns/tCr, tNAA/tCr, and tCho/tCr and the MR spectrum of lesions. Green arrow points toward a “rim” lesion, which is clearly visible on mIns/tNAA and tNAA/tCr.

**Figure 5:**
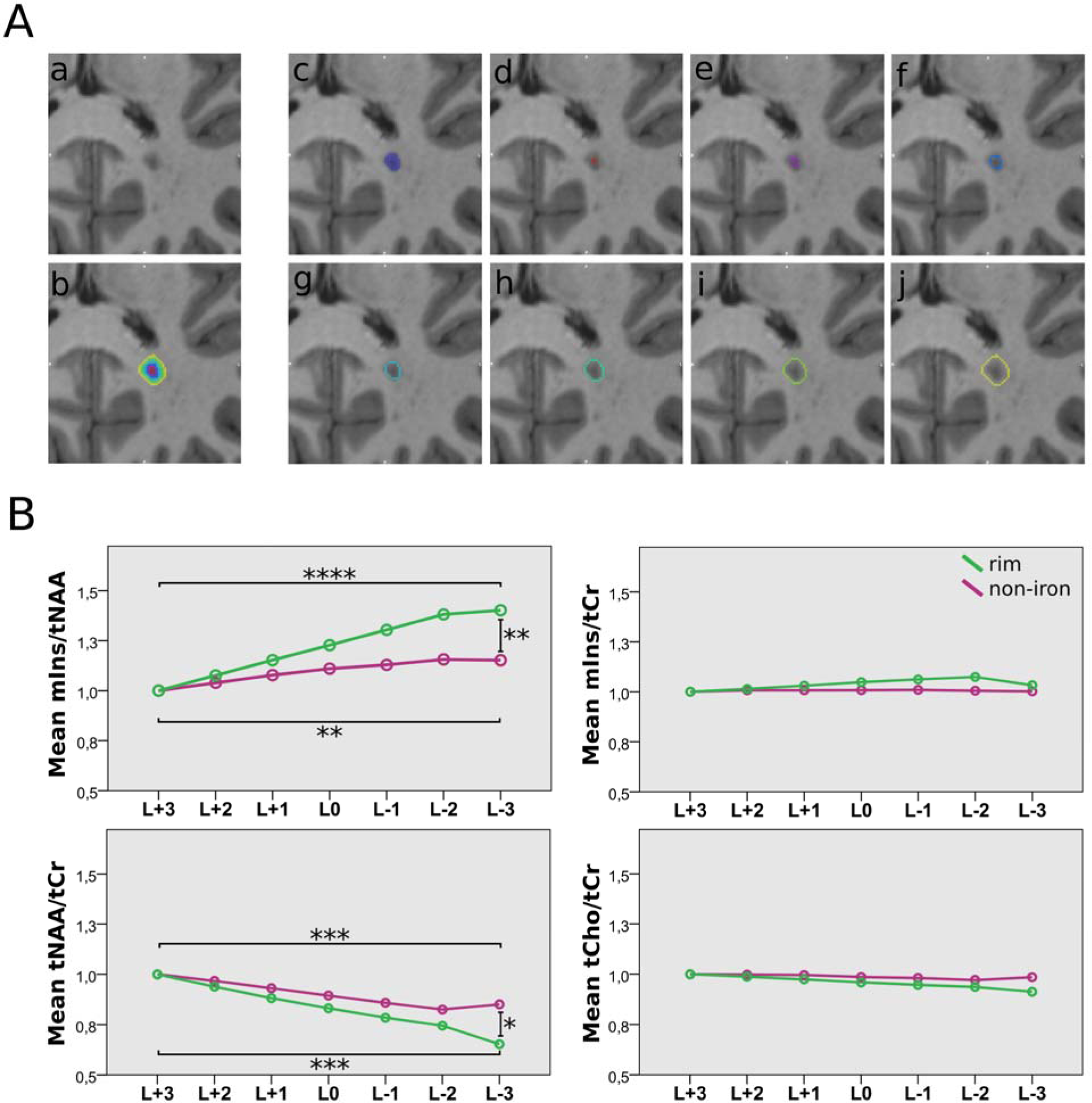
A: An example of an MS lesion (a) and seven lesion layer rings (d)-(j) created by dilation/erosion of the segmented mask (c) using mincmorph. Dilation of lesions close to the GM or cerebrospinal fluid (CSF) can potentially lead to a lesion layer ring intruding into the GM or CSF; thus, lesion-free GM and CSF masks were created using Freesurfer and mincmath. To cancel out intruding voxels, these lesion-free masks were subtracted from the dilated lesion layer rings. Manual corrections with FSLView were performed if necessary. (b) All layer rings merged. B: Boxplot diagram of normalized metabolic ratios show differences in mIns/tNAA and tNAA/tCr between the lesion layers of “rim” and “non-iron” lesions. Metabolic ratios of all layers were normalized to the outermost layer (L+3). Only significant results between L+3, L0, and L-3 are shown.

#### 2.4.3. Longitudinal analysis of newly emerging lesions

As the patient cohort overlapped with a previous longitudinal study (Dal-Bianco et al., 2021), morphological imaging data for additional time points were available. Thus, it was possible to identify newly emerging lesions and determine their age. These lesions—if spectroscopic data was available for the respective time point—were segmented and the metabolic patterns within each ROI were evaluated at all timepoints (i.e., prior and after appearance of the lesion on cMRI). The following time point ROIs were evaluated: “Year −1” (NAWM with the same position and size as the lesion occurring in Year 0); “Year 0” (newly identified lesion); and “Year 1” (follow-up measurement after one year). All these lesions were categorized according to their iron accumulation type (“non-iron” vs. “iron”).

#### 2.4.4. Statistics

Statistical analysis was performed using IBM SPSS 24. For all ROIs, descriptive statistics for multiple metabolic ratios (mIns/tNAA, Ins/tCr, tNAA/tCr, tCho/tCr) were derived. Using one-way ANOVA and Tukey post-hoc analysis (p<0.05 considered as statistically significant), metabolite levels between iron accumulation types, lesion layers (“rim” vs “non-iron” lesions), and newly emerging lesions (“Year −1” vs “Year 0” vs “Year 1” & “all new” vs “iron” vs “non-iron”) were compared. All values are listed as mean+-SD unless stated otherwise.

## 3. Results

Thirty-one patients with relapsing-remitting MS (16 female/15 male; mean age, 36.9 ± 10.3 years), were included in the final study. Patient demographics, including medications and EDSS scores (assessed in consensus by two experienced neurologists; A.DB, P.R), were collected (Table 1). In total, 487 ROIs were segmented: 412 “non-iron”; 31 “area;” eight “transition;” and 36 “rim.”.Of the 487 lesion ROIs, 220 fulfilled the minimum size criterion and were included in the final analysis (174 “non-iron,” 13 “area,” seven “transition,” and 26 “rim”). Forty-four “non-iron” and eight “rim” fulfilled the minimum size criterion for the layer analysis. For the newly emerging lesion analysis, 39 new lesions were identified in seven patients, with 27 having available spectroscopic data, resulting in 11 “non-iron” and 16 “iron” lesions analyzed (Table 2).

**Table 1:** Characteristics and Clinical Data of RRMS Patients.

| Characteristic | Participants with RRMS (n = 31) |
| --- | --- |
| Sex |  |
| Male | 15 |
| Female | 16 |
| Age (y) * | 36.9 ± 10.3 (21 - 55) |
| Disease Duration (months) * | 109.42 ± 61.94 (2 - 229) |
| EDSS score † | 1 (0 - 3.5) |
| No. of |  |
| MS lesions ° | 487 (220) |
| non-iron ° | 412 (174) |
| area ° | 31 (13) |
| transition ° | 8 (7) |
| rim ° | 36 (26) |
| No. of participants receiving therapy | 24 |
| first-line | 13 |
| second-line | 11 |

**Table 2:** Characteristics of data for the evaluation of newly emerging lesions.

| Characteristic | Participants with RRMS (n = 31) |
| --- | --- |
| No. of |  |
| patients with newly emerging lesions | 7 |
| patients receiving therapy* | 6 |
| first-line | 5 |
| second-line | 1 |
| newly emerging lesions† | 39 |
| non-iron | 21 |
| iron | 18 |
| lesions with spectroscopic data available | 27 |
| non-iron | 11 |
| iron | 16 |

\* no changes of treatment regimen within the investigated time period
† 2 of 39 newly emerging lesions found in the non-treated patient

The spectral quality was high and, even though NAA is reduced by pathology, the mean SNR of NAA ranged from 11-15, and the mean FWHM of NAA ranged from 19.6-21.4Hz, while the average CRLB values (mIns, tCho, tCr, tNAA) ranged from 15-26% (Suppl. Table 2).

### 3.1. Iron accumulation type comparison

Differences in mIns/tNAA were the most prominent metabolic distinction between different iron accumulation types (Table 3, Figure 3&4): “rim” lesions had, on average, 35% higher mIns/tNAA than “non-iron” (1.53±0.97 vs. 1.13±0.58, p=0.02) and 65% higher mIns/tNAA than “area” lesions (0.93±0.30, p=0.035). Furthermore, in both “non-iron” (1.13±0.58, p≤0.0001) and “rim” (1.53±0.97, p≤0.0001) lesions, mIns/tNAA was higher than in “NAWM” (0.53±0.17).

**Table 3:**
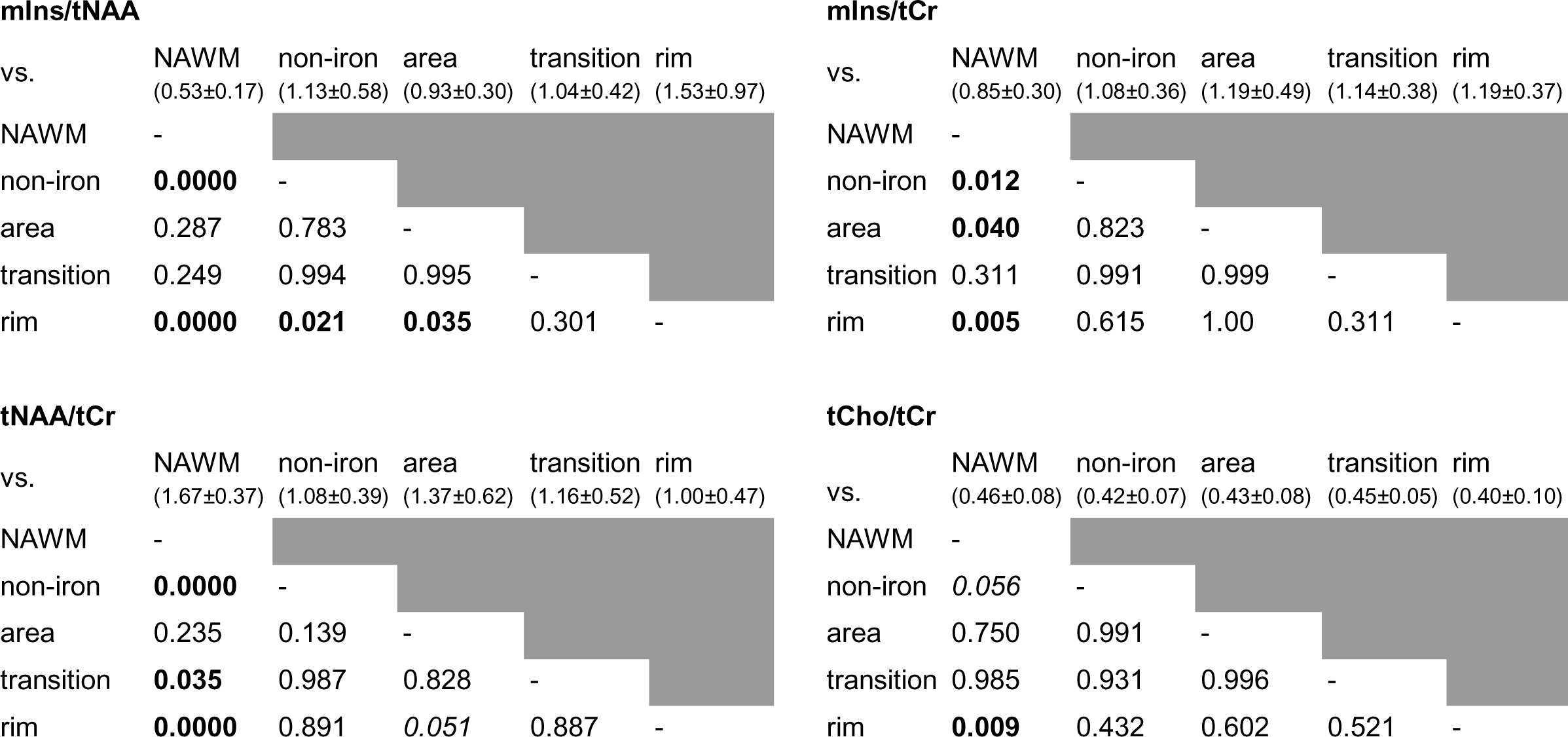
p-values of the Iron Accumulation Type Comparison. Mirrored cells (dark gray) were left out for better readability of the table. All metabolic concentration ratio values are listed as mean+-SD.

mIns/tNAA
| vs. | NAWM<br>(0.53±0.17) | non-iron<br>(1.13±0.58) | area<br>(0.93±0.30) | transition<br>(1.04±0.42) | rim<br>(1.53±0.97) |
| --- | --- | --- | --- | --- | --- |
| NAWM | - |  |  |  |  |
| non-iron | **0.0000** | - |  |  |  |
| area | 0.287 | 0.783 | - |  |  |
| transition | 0.249 | 0.994 | 0.995 | - |  |
| rim | **0.0000** | **0.021** | **0.035** | 0.301 | - |

mIns/tCr
| vs. | NAWM<br>(0.85±0.30) | non-iron<br>(1.08±0.36) | area<br>(1.19±0.49) | transition<br>(1.14±0.38) | rim<br>(1.19±0.37) |
| --- | --- | --- | --- | --- | --- |
| NAWM | - |  |  |  |  |
| non-iron | **0.012** | - |  |  |  |
| area | **0.040** | 0.823 | - |  |  |
| transition | 0.311 | 0.991 | 0.999 | - |  |
| rim | **0.005** | 0.615 | 1.00 | 0.311 | - |

tNAA/tCr
| vs. | NAWM<br>(1.67±0.37) | non-iron<br>(1.08±0.39) | area<br>(1.37±0.62) | transition<br>(1.16±0.52) | rim<br>(1.00±0.47) |
| --- | --- | --- | --- | --- | --- |
| NAWM | - |  |  |  |  |
| non-iron | **0.0000** | - |  |  |  |
| area | 0.235 | 0.139 | - |  |  |
| transition | **0.035** | 0.987 | 0.828 | - |  |
| rim | **0.0000** | 0.891 | 0.051 | 0.887 | - |

Table 3: p-values of the Iron Accumulation Type Comparison. Mirrored cells (dark gray) were left out for better readability of the table. All metabolic concentration ratio values are listed as mean ± SD.
| vs. | NAWM<br>(0.46±0.08) | non-iron<br>(0.42±0.07) | area<br>(0.43±0.08) | transition<br>(0.45±0.05) | rim<br>(0.40±0.10) |
| --- | --- | --- | --- | --- | --- |
| NAWM | - |  |  |  |  |
| non-iron | 0.056 | - |  |  |  |
| area | 0.750 | 0.991 | - |  |  |
| transition | 0.985 | 0.931 | 0.996 | - |  |
| rim | **0.009** | 0.432 | 0.602 | 0.521 | - |

For mIns/tCr, no differences were found between “rim” (1.19±0.37) lesions and other lesion types (vs. “non-iron” p=0.615; vs. “area” p=1.00), but mlns/tCr was higher than that in “NAWM” (0.85±0.30, p=0.005). mIns/tCr in “NAWM” was also lower compared to “non-iron” (1.08±0.36, p=0.012) and “area” (1.19±0.49, p=0.040) lesions. “Area” was not different from “non-iron” (p=0.823) and “transition” (1.14±0.38) did not show any differences (vs. “NAWM” p=0.311; vs. “non-iron” p=0.991; vs. “area” p=0.999; vs. “rim” p=0.998).

In “NAWM,” the tNAA/tCr (1.67±0.37) was higher than in “non-iron” (1.08±0.39, p≤0.001), “rim” (1.00±0.47, p≤0.001), and “transition” (1.14±0.38, p=0.035) lesions. There were no differences in NAA/tCr between iron-deposition categories, although there was a trend toward higher NAA/tCr in “rim” vs. “area” lesions (1.19±0.49,p=0.051).

Compared to “NAWM” (0.46±0.08), there was lower tCho/tCr in “rim” (0.40±0.10, p=0.009) lesions; however this was not the case in “non-iron” (0.42±0.07, p=0.056) lesions.

### 3.2. Lesion periphery

All reported results (Table 4, Figure 5B) were achieved through normalization using the outermost layer, thus resulting in all L+3 layers having a value of 1.00±0.00 and no comparisons of L+3 between “iron” and “non-iron” being considered.

**Table 4:**
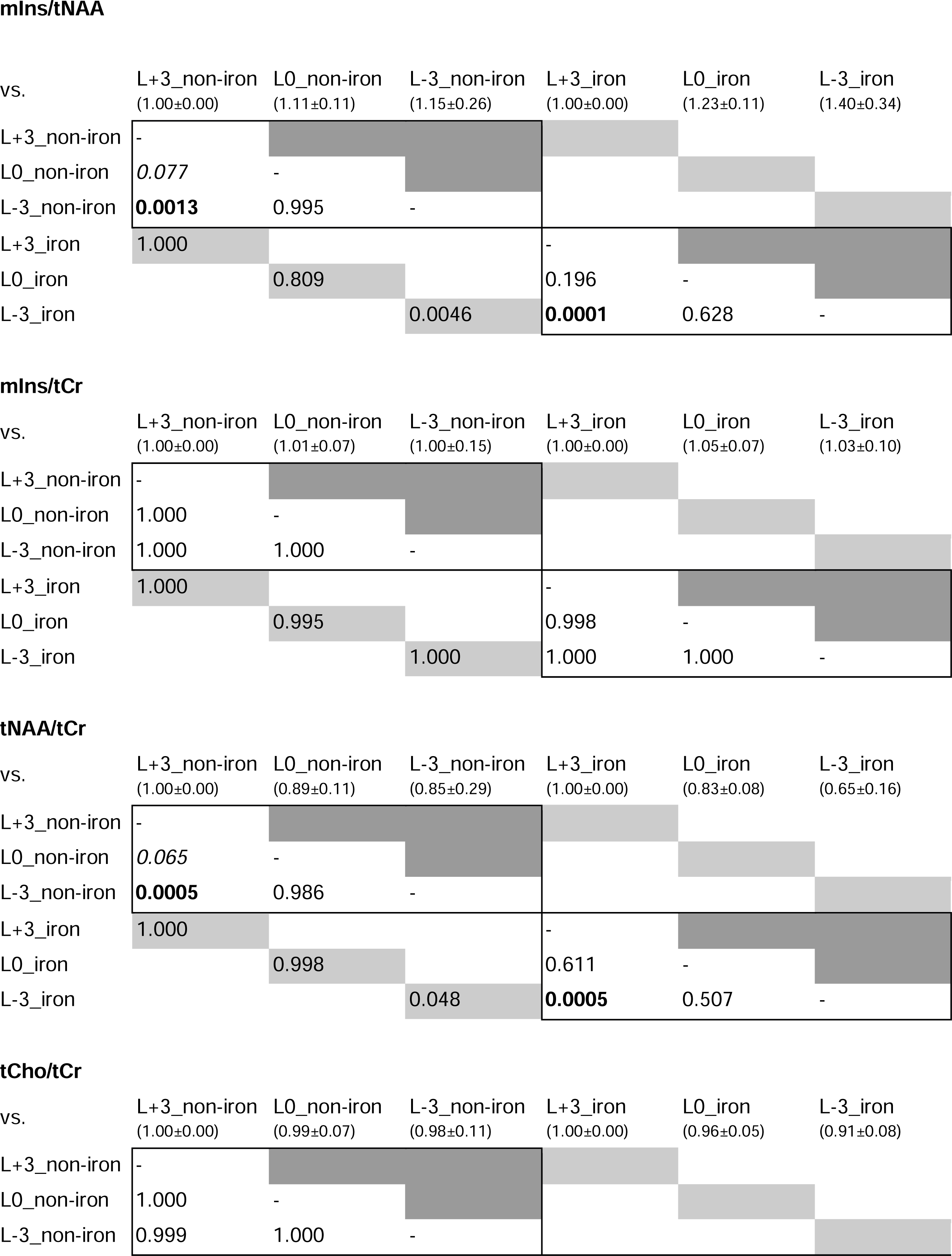
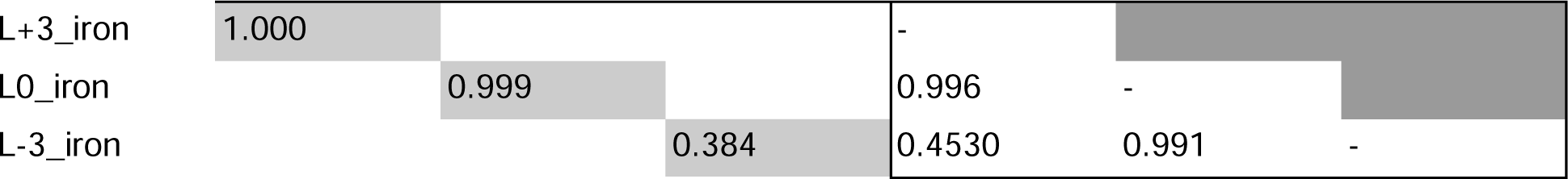
p-values of the Lesion Periphery Analysis. Mirrored cells (dark gray) were left out for better readability of the table. Light gray cells reflect comparisons between iron types within the respective layer, which are depicted only once for better readability. All metabolic concentration ratio values are listed as mean+-SD.

Differences between lesion layers within and between iron accumulation types were found for mIns/tNAA and tNAA/tCr between layers L-3 and L+3.

In the “non-iron” lesions subgroup, there was a 15.2% higher mIns/tNAA in the innermost layer (L-3) than in the outermost layer (L+3) (1.15±0.26 vs. 1.00±0.00, p≤0.01). In contrast, the group of “rim” lesions had a 40.22% higher mIns/tNAA in the L-3 compared to the L+3 layer (1.40±0.34 vs. 1.00±0.00, p≤0.001).

Between the groups (“rim” vs. “non-iron”), “rim” exhibited a 21.71% higher mIns/tNAA for L-3 compared to “non-iron” (mean±SD: 1.40±0.34 vs. 1.15±0.26, p=0.005).

For tNAA/tCr, an inverse behavior was observed. In the “non-iron” group, L-3 had a −14.94% lower tNAA/tCr than L+3 (0.85±0.29 vs. 1.00±0.00, p≤0.001). In the “rim” group, L-3 exhibited a −34.73% lower tNAA/tCr compared to L+3 (0.65±0.16 vs. 1.00±0.00, p≤0.001). Between the groups, tNAA/tCr of the L-3 layer was −23.27% lower in “rim” than in “non-iron” lesions (0.65±0.16 vs. 0.85±0.29, p=0.048)

For both mIns/tCr and tCho/tCr, no differences within or between the groups were significant.

### 3.3. Newly emerging lesions

Significant differences in newly emerging lesions were found for mIns/tNAA and tNAA/tCr (Table 5, Figure 6).

**Figure 6:**
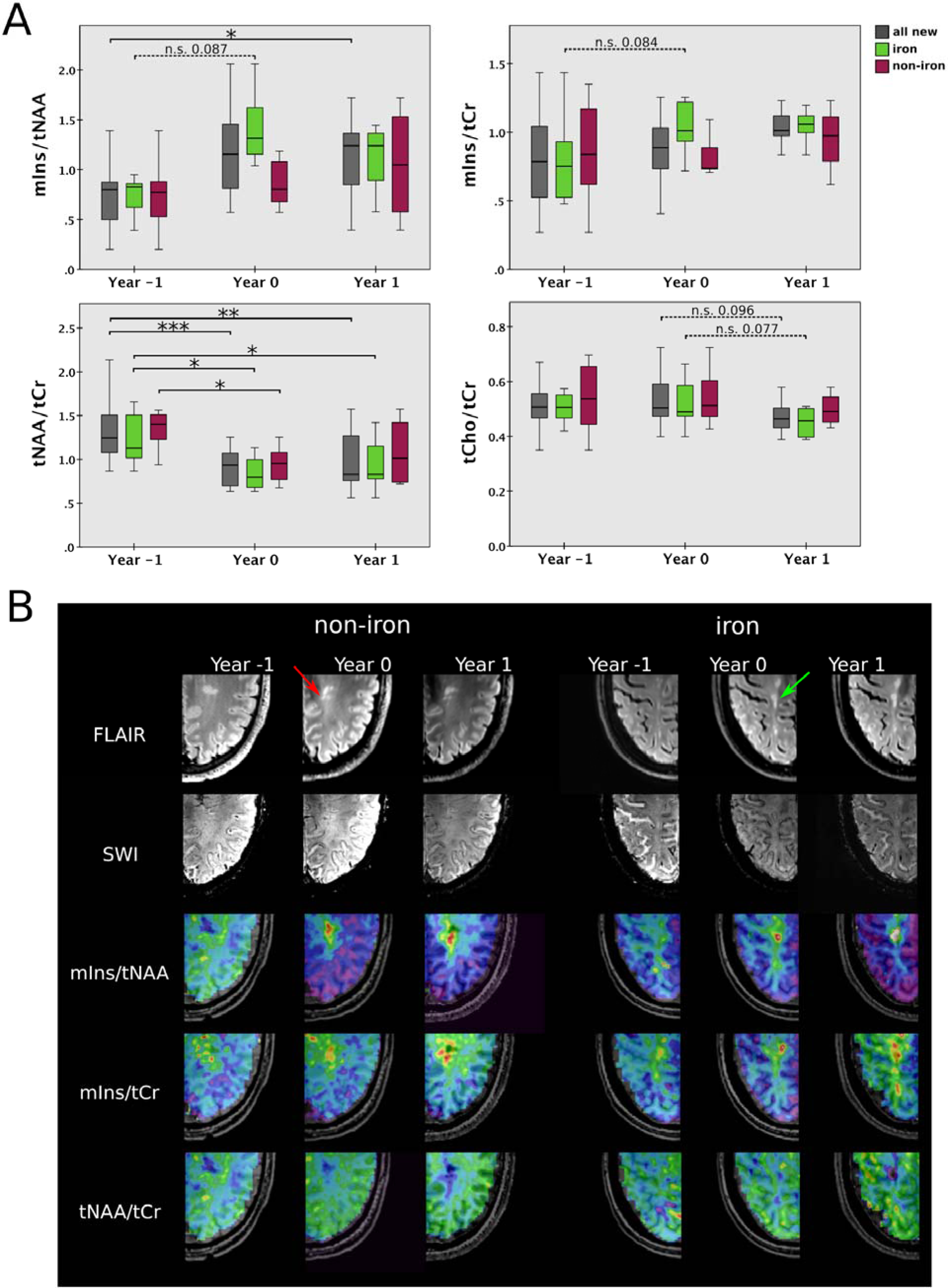
A: Boxplot diagram for mIns/tNAA, mIns/tCr, tNAA/tCr, and tCho/tCr of newly emerging “non-iron” and “iron” lesions (also of joint “all new” group) for “Year −1” “Year 0,” and “Year 1.” Significant results were found especially for tNAA/tCr. B: T2-weighted FLAIR and SWI, as well as metabolic maps of mIns/tNAA, mIns/tCr and tNAA/tCr for an exemplary “non-iron” and “iron” lesion at the yearly follow-up (“Year −1,” “Year 0,” and “Year 1”).

**Table 5:**
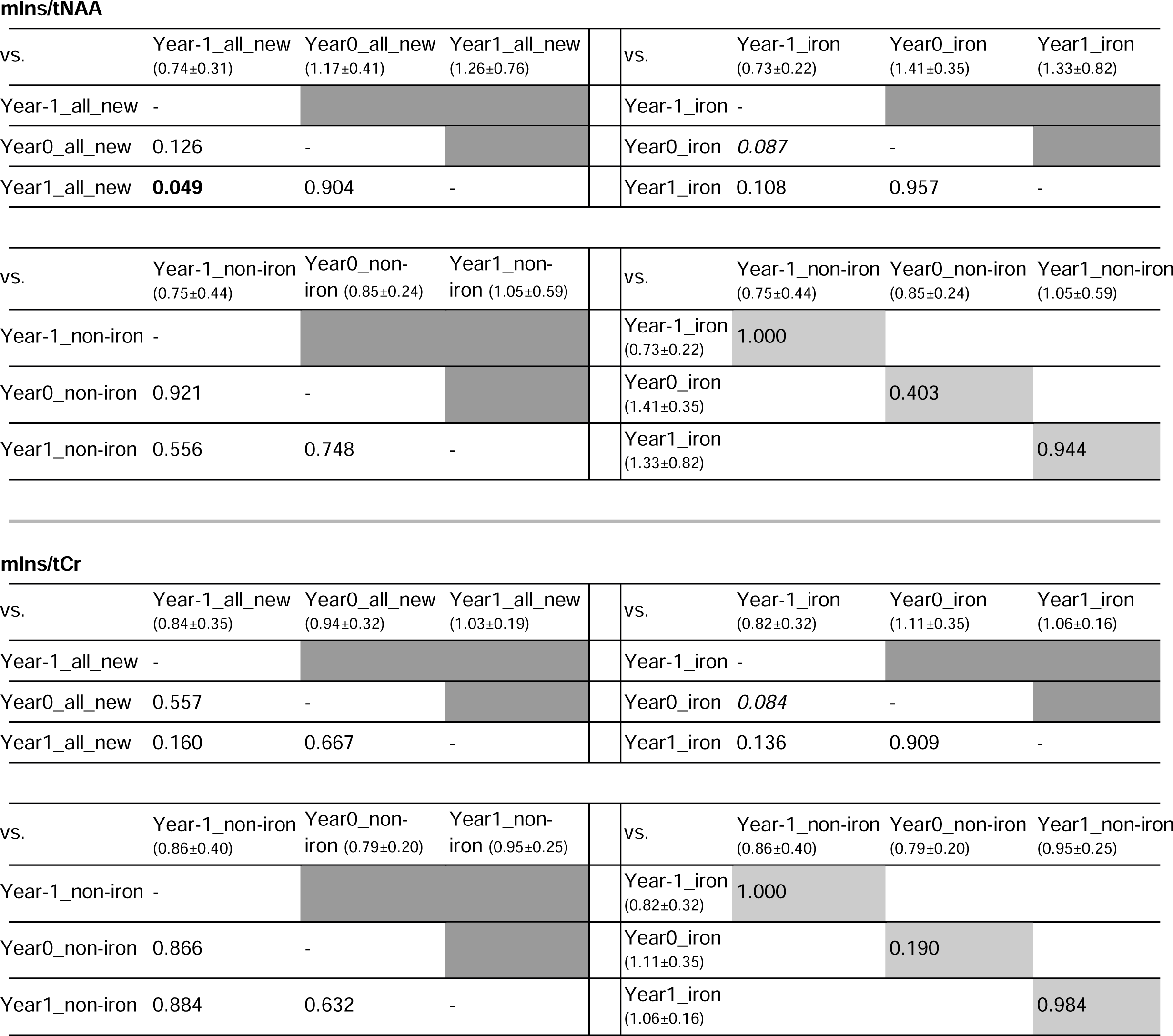
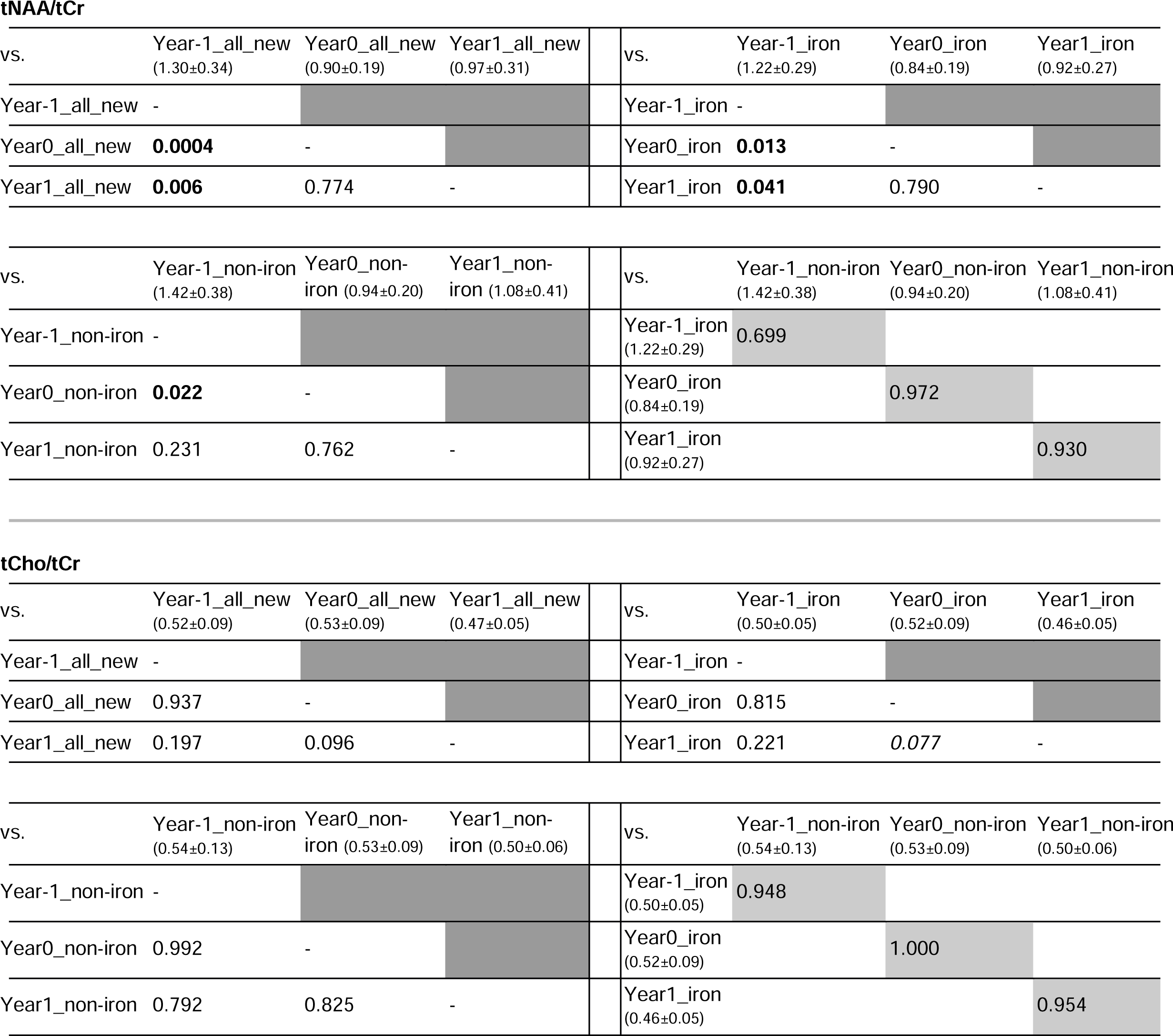
p-values of the Newly Emerging Lesions Analysis. Mirrored cells (dark gray) were left out for better readability of the table. Light gray cells reflect comparisons between iron types within the respective year. All metabolic concentration ratio values are listed as mean+-SD.

A 69.6% increase of mIns/tNAA was observed in newly identified lesions between “Year −1” and “Year 1” (0.74±0.31 vs. 1.26±0.76, p=0.049). There was no difference in mIns/tNAA between “Year −1” and “Year 0” (0.74±0.31 vs. 1.17±0.41, p=0.126) or between “Year 0” and “Year 1” (1.17±0.41 vs. 1.26±0.76, p=0.904).

No differences were found in mIns/tCr during lesion development.

The most significant changes during lesion development were found for tNAA/tCr: in the group of all new lesions, “Year 0” (0.90±0.20, p≤ 0.001) and “Year 1” (0.97±0.31, p=0.006) were lower by −31% and −26%, respectively, compared to “Year −1” (1.30±0.34). In “iron” lesions, tNAA/tCr was lower by −31% and −24%, respectively, in “Year 0” (0.84±0.19, p=0.013) and “Year 1” (0.92±0.27, p=0.041) when compared to “Year −1” (1.21±0.29). In “non-iron” lesions, tNAA/tCr of “Year 0” was −34% lower than that of “Year −1”(1.42±0.38 vs. 0.95±0.20, p=0.022), while there were no significant differences between “Year −1” vs. “Year 1” (1.08±0.41, p=0.231).

For tCho/tCr, no significant results were found.

## 4. Discussion

This study evaluated the frequency of iron deposition types and the occurrence of iron accumulation in newly emerging lesions in 31 RRMS patients at 7T. It further evaluated metabolic alterations within these different iron deposition types, inside and in the proximity of (non)-iron-containing lesions and the longitudinal metabolic changes during the early development of lesions according to their iron status.

We found that less than half (48%) of iron-containing lesions showed a distinct paramagnetic rim of iron accumulation and that 46% of lesions already showed iron accumulation when newly emerging. Furthermore, MS lesions were clearly visualized on high-resolution metabolic maps, especially those of tNAA/tCr and mIns/tNAA.

In general, all MS lesions showed metabolic differences compared to NAWM. The metabolic changes between non-iron and iron-containing lesions were driven by both tNAA and mIns, while most axonal damage was found in iron-containing lesions with a distinct rim (Tozlu et al., 2021). Looking into the other iron accumulation subgroups, the “transition” type is both macroscopically and metabolically an intermediate between iron rim lesions and iron-containing lesions with a diffuse iron accumulation. These “area” lesions interestingly showed a 38% higher tNAA/tCr compared to iron rim lesions and a 27% higher tNAA/tCr compared to non-iron lesions, possibly representing an intrinsic attempt to enhance neural integrity, supporting the presumed role of iron as a co-regulator of remyelination (Stephenson et al., 2014).

Unfortunately, the number of “area” lesions that met the minimum volume criterion was low. Thus, only 42% could be evaluated in the final analysis, resulting in an underpowered, and thus, non-significant statistical comparison to rim lesions. The finding of increased tNAA/tCr in “area” lesions is in agreement with a previous preliminary study (Lipka et al., n.d.). Nevertheless, these previous preliminary results, as well as the frequency of iron-containing lesions without a distinct rim and their different metabolic profiles, highlights the importance of characterizing iron-containing lesions into distinct groups (Hametner et al., 2018), as different types of iron accumulation might have a different influence on disease course.

We also investigated metabolic profiles inside and at the periphery of lesions. A steeper metabolic gradient (i.e., higher mIns/tNAA and lower tNAA/Cr in the center) was found for rim lesions compared to non-iron lesions. We found that tNAA reduction is the predominant differentiator between non-iron and rim lesions. This reconfirms histological studies (Absinta et al., 2016; Maggi et al., 2021) that reported more damage inside iron rim lesions and the role of tNAA as a biomarker for tissue damage (Lipka et al., 2023), explaining the worse clinical outcome (Absinta et al., 2019) in patients with distinct rim-shaped iron-accumulating lesions. Nevertheless, we did not find any difference between non-iron and rim lesions with regard to mIns (Lipka et al., 2023), which would have confirmed the importance of iron rims in the ongoing debate on slowly expanding lesions (Absinta & Dal-Bianco, 2021; Arnold et al., 2021; Enzinger, 2021).

We found reduced tNAA in newly emerging lesions regardless of iron accumulation, confirming results by Kirov et al. (Kirov et al., 2017) in persistent lesions, although their study incorporated only three lesions and did not differentiate between iron accumulation types. Looking into our one-year follow-up scans, tNAA showed a partial re-elevation in non-iron lesions, while it remained reduced in iron-containing lesions, similar to the results of Kirov et al. (Kirov et al., 2017) for resolving lesions, although their study was performed at 3T using a much lower spatial resolution, while having substantially longer measurement times and only including 10 patients. Our results, therefore, reconfirm the higher damage in iron-containing lesions and their reduced remyelination, although for this analysisLdue to the low number of newly emerging lesionsLno difference between iron accumulation types was found. Furthermore, the longitudinal assessment of mIns in our study suggests that mIns might have a larger influence on earlier lesion development in iron-containing lesions compared to non-iron lesions, potentially reflecting a higher accumulation of reactive astroglia. Even though not many newly emerging lesions were found due to the patient population having a stable disease course, almost half the lesions did show iron accumulation from the beginning, which might be the determining factor in disease progression (Absinta et al., 2016; Treaba et al., 2021; Altokhis et al., 2022; Harrison et al., 2016; Maggi et al., 2021; Marcille et al., 2022).

In addition to the already mentioned limited sample size of “area” and newly emerging lesions, our study has some further limitations. As our MRSI was limited to a single 8-mm thick slice above the corpus callosum the overall number of included lesions was limited. Additionally, the lesion layer thickness was restricted, thereby resulting in a moving average. Moreover, the number of included lesions had to be further reduced in order to avoid bias due to partial volume effects in z-direction. Thereby, in-plane partial volume errors were minimized, and large pathological changes as seen in MS lesions still prevail. For the analysis of lesion layers the influence of partial volume errors could not be fully extinguished, even though only every third lesion layer has been analyzed. To overcome these limitations of a single-slice acquisition, a 3D-MRSI version of this sequence, covering the majority of the brain, has recently been introduced and has provided promising results in brain tumors (Hangel et al., 2022; Hingerl et al., 2020). We did not determine concentration estimates (Kreis et al., 2021; Maudsley et al., 2021), as we could not avert changes in the T1 relaxation times due to MS pathology (Brief et al., 2010) and an additional water scan would have doubled our acquisition time (Kreis et al., 2021; Maudsley et al., 2021). Changed levels in metabolic ratios, e.g., an increased level of mIns/tNAA can result from either an increased nominator, a decreased denominator or a combined effect. Using the ratio of mIns and tNAA, each individually showing changes of high magnitude on single metabolic maps, had a multiplying effect on the clear distinction of metabolic alterations. Single metabolite maps of tCr did not show any metabolic changes due to pathology. Therefore it was chosen as a reference to study mIns, tNAA and tCho. Even though this might have led to an underestimation of respective metabolic alterations, these changes still remain clearly distinguishable. Moreover, it must be taken into account that in vivo characterization of the inflammatory activation stage during lesion development is still limited. The dynamics of lesion development progresses from a highly active to a chronic active to an inactive, remyelinating stage. The highly active lesions can be readily visualized with contrast agent (Gonzalez-Scarano et al., 1987; Absinta et al., 2013), whereas a subtype of chronic active lesions are characterized by iron rims (Absinta et al., 2016; Dal-Bianco et al., 2017, 2021; Weber et al., 2022), but distinct identification of inactive and remyelinating lesions is still lacking. Here, long-term observations of newly emerging lesions (ideally with a shorter interval of e.g. 6 months) may be able to provide promising molecular insight. Thus, MR spectroscopy might provide an important contribution to the characterization of the lesion type based on the molecular changes in and around the lesion.

## 5. Conclusion

In this study, we observed metabolic alterations that were associated with iron deposition in MS lesions. In particular, we found lower tNAA in the center of iron-containing lesions with a distinct rim compared to non-iron lesions, reflecting more severe tissue damage. Furthermore, in newly emerging lesions with iron-accumulation, the tNAA decrease was persistent on a follow-up scan in contrast to non-iron lesions, where tNAA partially recovered on a follow-up scan. Future studies should, therefore, take different types of iron accumulation into account, as each type is likely associated with a different level of tissue damage, as indicated by their different metabolic profiles.

## Abbreviations

EDSS: Expanded Disability Status Scale
mIns: myo-inositol
tNAA: total N-acetyl aspartate
FID-MRSI: free-induction decay magnetic resonance spectroscopic imaging
MS: multiple sclerosis
NAWM: normal-appearing white matter
SWI: Susceptibility Weighted Imaging
tCr: total creatine
tCho: total choline

## Data Availability

All data produced in the present study are available upon reasonable request to the authors

