## Supplementary Table 1, Supplementary Table 2 & Supplementary Text 1 for "Metabolic Insights into Iron Deposition in Relapsing-Remitting Multiple Sclerosis via 7T Magnetic Resonance Spectroscopic Imaging"

| **MRSinMRS checklist** | |
| --- | --- |
| 1. Hardware |  |
| a. Field strength [T] | 7 |
| b. Manufacturer | Siemens |
| c. Model (software version if available) | Magnetom 7T |
| d. RF coils: nuclei (transmit/ receive), number of channels, type, body part | 1H, 32 ch, head, Nova Medical |
| e. Additional hardware | N/A |
| 2. Acquisition |  |
| a. Pulse sequence | FID-MRSI |
| b. Volume of Interest (VOI) locations | Parallel to the anterior commissure-posterior commissure line, superior to the lateral ventricles, and covering the centrum semiovale region |
| c. Nominal VOI size [cm3, mm3 ] | 220×220×20 mm3 |
| d. Repetition Time (TR), Echo Time (TE) [ms, s] | TR=200 ms / 1.3 ms acquisition delay |
| e. Total number of Excitations or acquisitions per spectrum | 1 average |
| In time series for kinetic studies | N/A |
| i. Number of Averaged spectra (NA) per time-point | N/A |
| ii. Averaging method (e.g. block-wise or moving average) | N/A |
| iii. Total number of spectra (acquired / in time-series) | N/A |
| f. Additional sequence parameters (spectral width in Hz, number of spectral points, frequency offsets); If STEAM: Mixing Time TM; If MRSI: 2D or 3D, FOV in all directions, matrix size, acceleration factors | BW 6000 Hz, 1024 spectral points, MRSI: 2D, FOV 220×220 mm2 , 8mm slice thickness, matrix size 100×100, Acc. factor: 4 |
| g. Water Suppression Method | WET |
| h. Shimming Method, reference peak, and thresholds for “acceptance of shim” chosen | Standard shim + manual adjustment, water peak < 40 Hz |
| i. Triggering or motion correction method (respiratory, peripheral, cardiac triggering, incl. device used and delays) | N/A |
| 3. Data analysis methods and outputs |  |
| a. Analysis software | LCModel 6.3-1 |
| b. Processing steps deviating from quoted reference or product | N/A |
| c. Output measure (e.g. absolute concentration, institutional units, ratio) | institutional units, ratio |
| d. Quantification references and assumptions, fitting model assumptions | Simulated in NMRScope-B, macromolecular background |
| 4. Data Quality |  |
| a. Reported variables (SNR, Linewidth (with reference peaks)) | SNR was calculated using the pseudoreplica method, and linewidth as FWHM of the NAA fit (see Supplementary Table 2) |
| b. Data exclusion criteria | Volume below 20mm³ for all lesions |
| c. Quality measures of postprocessing Model fitting (e.g. CRLB, goodness of fit, SD of residual) | CRLB (see Supplementary Table 2) |
| d. Sample Spectrum | See Figure 4 |

**Supplementary Table 1:** Minimum Reporting Standards for ***in vivo*** MR Spectroscopy

|  | CRLB_mIns_ [%] | CRLB_tCho_ [%] | CRLB_tCr_ [%] | CRLB_tNAA_ [%] | SNR | FWHM_NAA_ [Hz] |
| --- | --- | --- | --- | --- | --- | --- |
| NAWM | 25.93 ± 5.99 | 16.42 ± 3.68 | 19.18 ± 3.45 | 15.31 ± 3.08 | 15.33 ± 3.01 | 20.44 ± 2.83 |
| non-iron lesions | 21.96 ± 7.15 | 18.42 ± 7.07 | 20.33 ± 6.48 | 21.21 ± 7.45 | 11.49 ± 4.30 | 21.44 ± 3.73 |
| iron lesions | 21.75 ± 7.00 | 19.34 ± 9.40 | 19.98 ± 8.15 | 22.18 ± 7.93 | 11.17 ± 4.84 | 19.64 ± 2.73 |

**Supplementary Table 2:** Spectral and fitting quality metrics for each investigated metabolite and study group.

Lipka, A., Niess, E., Dal-Bianco, A., Hangel, G. J., Rommer, P. S., Strasser, B., Motyka, S., Hingerl, L., Berger, T., Hnilicová, P., Kantorová, E., Leutmezer, F., Kurča, E., Gruber, S., Trattnig, S., & Bogner, W. (2023). Lesion-Specific Metabolic Alterations in Relapsing-Remitting Multiple Sclerosis Via 7 T Magnetic Resonance Spectroscopic Imaging. *Investigative Radiology*, *58*(2), 156. https://doi.org/10.1097/RLI.0000000000000913

→ of the 51 RRMS patients studied in the above mentioned study, 31 RRMS patients were also included in this study. In the previous study the focus was on focal metabolic alterations inside and in the periphery of lesions that are visible or invisible on conventional MRI and to correlate metabolic changes with T1-hypointensity and distance between lesions and cortical gray matter. In contrast to that, the current study reports on SWI data and correlates the information on lesion iron accumulation to MRSI in multiple aspects.

A. Dal-Bianco et al., “Long-term evolution of multiple sclerosis iron rim lesions in 7 T MRI,” Brain, vol.

144, no. 3, pp. 833–847, Mar. 2021, doi: 10.1093/brain/awaa436

→ the above mentioned study included 33 patients (30 RRMS, 3 SPMS) in a longitudinal study

(longest follow up duration of 7 years in 8 patients). The 30 RRMS patients were partly involved in

the study, if they did not drop out until 2017. Furthermore, the study did only focus on imaging

features such as lesion volume and T1 and assessed these features longitudinally within the RRMS

and SPMS cohort. The study did not report on any MRSI data.

**Supplementary Text 1:** Overlapping study statement
